# Sit-up Test for Assessing Blood Pressure Dysregulation in Community-Dwelling Older Adults: A Cross-Sectional Study

**DOI:** 10.1101/2025.02.05.25321761

**Authors:** Kazuaki Oyake, Yoshiharu Yokokawa

## Abstract

**Background:** The sit-up test enables the assessment of orthostatic hypotension without using a tilt table in individuals at high risk of falling when standing; however, no studies have compared blood pressure responses between those with and without orthostatic hypotension during this test. The primary objective of this study was to compare blood pressure responses during the sit-up test between community-dwelling older adults with and without orthostatic hypotension. The secondary objective was to determine the associations between orthostatic hypotension detected by the sit-up test and poor health conditions in these individuals.

**Methods:** One hundred-two community-dwelling older adults underwent the sit-up test. Orthostatic hypotension was defined as a decrease of ≥10 mmHg in systolic blood pressure and/or ≥5 mmHg in diastolic blood pressure during the test. Supine and seated hypertension were evaluated, defined as systolic blood pressure ≥ 140 mmHg and/or diastolic blood pressure ≥ 90 mmHg. Blood pressure responses during the test were compared between participants with and without orthostatic hypotension. Moreover, associations of orthostatic hypotension with demographic, clinical, and geriatric outcomes were examined.

**Results:** Thirty-four participants (33.3%) showed orthostatic hypotension during the test. Participants with orthostatic hypotension demonstrated a greater decrease in systolic blood pressure (F_(3,297)_ = 47.0, p < 0.001), a smaller increase in diastolic blood pressure (F_(3,297)_ = 26.5, p < 0.001), and higher supine systolic blood pressure (t = 3.363, p = 0.005) during the test than those without orthostatic hypotension. Consequently, 52.9% of participants with orthostatic hypotension had supine hypertension. Orthostatic hypotension was associated with a higher proportion of participants with at least one comorbidity (odds ratio = 4.50, p = 0.002) and those with non-robust status (odds ratio = 3.08, p = 0.022), even after adjusting for supine and seated hypertension.

**Conclusion:** Community-dwelling older adults with orthostatic hypotension were characterized by an impaired orthostatic increase in diastolic blood pressure and high supine systolic blood pressure during the sit-up test. Orthostatic hypotension was associated with poor health conditions, independently of supine and seated hypertension. These findings contribute valuable insights for the application of the sit-up test in preventive health screenings for older adults.

## Background

Blood pressure management in older adults requires careful consideration of the various physiological changes associated with aging. Hypertension, defined as systolic blood pressure ≥ 140 mmHg and/or diastolic blood pressure ≥ 90 mmHg in the sitting position (seated hypertension), is a major risk factor for mortality and cardiovascular diseases [1]. These relationships are partly mediated by arterial stiffness, which reduces baroreflex sensitivity [2]. Conventional seated blood pressure measurement remains the standard practice in routine health screenings; however, this single-position assessment may not fully capture blood pressure dysregulation. Reduced baroreflex sensitivity with aging may result in orthostatic hypotension and supine hypertension, in addition to seated hypertension [2].

Orthostatic hypotension, defined as a decrease in systolic blood pressure of ≥ 20 mmHg or diastolic blood pressure of ≥ 10 mmHg within 3 min of standing or head-up tilt, is a common manifestation of blood pressure dysregulation that is associated with increased risks of mortality and morbidity, including falls, dementia, cardiovascular diseases, and stroke, independent of hypertension [3–10]. The prevalence of orthostatic hypotension increases with aging, affecting 22.2% (95% confidence interval = 17–28) of community-dwelling older adults [11]. Moreover, approximately 50% of individuals with orthostatic hypotension have supine hypertension, defined as a systolic blood pressure of ≥ 140 mmHg and/or diastolic blood pressure of ≥ 90 mmHg in the supine position [12–14]. Supine hypertension is reportedly associated with a high risk of heart failure, stroke, and all-cause mortality, regardless of orthostatic hypotension status [14].

The sit-up test is developed to assess orthostatic hypotension in individuals who cannot independently stand or are at a high risk of falling when standing [15–18]. Orthostatic decreases in blood pressure elicited during sitting up are smaller than those during standing up owing to reduced acute changes in gravitational stress; therefore, the optimal cutoff points for orthostatic hypotension using the sit-up test are a decrease of 10 mmHg in systolic blood pressure or 5 mmHg in diastolic blood pressure [18]. In the sit-up test, participants are passively moved from the supine to the sitting position with the assessor’s assistance. This enables the assessment of three types of blood pressure dysregulation: orthostatic hypotension, supine hypertension, and seated hypotension.

Previous studies have examined hemodynamic responses to the sit-up test in older adults [19, 20]; however, to the best of our knowledge, no studies have compared blood pressure responses between those with and without orthostatic hypotension during this test. Additionally, no studies have investigated the associations between orthostatic hypotension detected by the sit-up test and adverse health outcomes in this population. Understanding these factors may enable the effective utilization of the sit-up test for the early identification and management of health deterioration in older adults. Therefore, the primary objective of this study was to compare blood pressure responses during the sit-up test between community-dwelling older adults with and without orthostatic hypotension. We hypothesized that participants with orthostatic hypotension would demonstrate higher supine systolic and diastolic blood pressure than those without orthostatic hypotension, considering that individuals with orthostatic hypotension frequently have supine hypertension [12–14]. Our secondary objective was to determine the associations between orthostatic hypotension detected by the sit-up test and poor health conditions in these individuals. We hypothesized that orthostatic hypotension would be associated with adverse health outcomes independently of seated hypertension.

## Methods

### Study design

This was a cross-sectional study. This study protocol was approved by the appropriate ethics committee of Shinshu University (approval number: 6281). All participants provided written informed consent before enrolment. We followed the Strengthening the Reporting of Observational Studies in Epidemiology reporting guidelines [21]. The study was performed in accordance with the 1964 Declaration of Helsinki, as revised in 2013.

We used Claude 3.5 Sonnet (Anthropic, San Francisco, CA, USA) for generating preliminary drafts and English editing assistance during the preparation of this work. We reviewed and edited the content after using this tool, and we assume full responsibility for the content of this publication.

### Participants

Participants were recruited from attendees of community-based health promotion classes held in the Shiga ward of Matsumoto City, Nagano, Japan. These classes, organized by the Community Development Division of Matsumoto City, were conducted at 27 different community centers between April 2023 and February 2024. Flyers with information on the classes were distributed to all households in the Shiga ward to publicize the study. Residents voluntarily participated in the classes. The inclusion criteria were participants (1) aged ≥ 65 years and (2) able to walk independently with or without assistive devices. Individuals were excluded if they had cognitive impairment or hearing loss preventing them from following the researcher’s instructions or they declined to participate in the sit-up testing.

### Assessments of demographic and clinical outcomes

The self-reported questionnaire included age, sex, height, and weight as demographic outcomes. A body mass index of < 18.5 kg/m^2^ was defined as being underweight, whereas that of ≥ 25.0 kg/m^2^ was defined as having obesity [22].

Additionally, the questionnaire included information on clinical outcomes, such as the number of prescribed medications, history of falls within a year, and comorbidities. Polypharmacy was defined as having ≥ 5 regular medications prescribed, excluding supplements [23]. A fall was described as an event resulting in a person unintentionally coming to rest on the ground or other lower-level surfaces [24]. Comorbidities included articular diseases, cardiac diseases, cancer, diabetes mellitus, respiratory diseases, and stroke. These diseases were typed out on the survey, and the participants selected all that applied to them.

### Assessment of geriatric outcomes

We assessed physical frailty and advanced glycation end products (AGEs) as geriatric outcomes. These outcomes have been associated with increased risks of adverse health outcomes, such as mortality and cardiovascular diseases [25–28]. Physical frailty is defined as a clinical syndrome of increased vulnerability owing to diminished strength, endurance, and physiological function, resulting in increased dependency [29]. We assessed physical frailty using the revised Japanese version of the Cardiovascular Health Study criteria, constituting five components: shrinking, exhaustion, low activity, slow gait speed, and weak handgrip strength [30]. Shrinking was defined as answering “yes” to the question “Have you unintentionally lost 2.0 kg or more in the past 6 months?” Exhaustion was defined as answering “yes” to “In the past 2 weeks, have you felt tired without a reason?” Low activity was defined by answering “no” to two questions: “Do you engage in moderate levels of physical exercise or sports aimed at health?” and “Do you engage in low levels of physical exercise aimed at health?” We measured the time required to walk 5 m at a comfortable speed [31]. Slow gait speed was defined as a comfortable gait speed of < 1.0 m/s. Handgrip strength was measured twice in the dominant hand, with the participant squeezing a Smedley-type hand grip dynamometer (T.K.K. 5401; SANKA Co., Ltd., Niigata, Japan) as hard as possible. The greater value of the two measurements was analyzed [31]. Handgrip strength < 28.0 kg for male participants and < 18.0 kg for female participants were considered weak handgrip strength. Participants were classified frail (≥ 3), pre-frail (1–2), or robust (none), based on the total number of positive items. We combined the pre-frail and frail groups into a non-robust group.

AGEs are a group of molecules generated nonenzymatically by sugars binding to proteins, lipids, or nucleic acids, resulting in protein modification and cross- linking [32]. AGE accumulation in tissues has been associated with age-related diseases, including diabetes, cardiovascular diseases, dementia, frailty, and sarcopenia [33–36]. AGEs can be non-invasively measured in the skin using skin autofluorescence [37]. Skin autofluorescence was measured from inside the forearm using a non-invasive device (AGE Reaser mu; DiagnOptics Technologies, Groningen, the Netherlands), which has been validated as a reliable and valid instrument [37]. Skin autofluorescence was quantified as the ratio of average autofluorescence per nanometer (nm) within the 420–600 nm range to the average autofluorescence per nm within the 300–420 nm range, measured over a 1 cm^2^ skin area. Skin autofluorescence values were expressed in arbitrary units. We ensured that the studied site lacked scars and had no cream applied. Participants performed the measurements in the sitting position, with the volar side of the forearm placed on top of the AGE reader. The mean of three consecutive measurements was used to avoid erroneous measurements.

### Sit-up test

The sit-up test was performed between 2:00 and 4:00 p.m. and more than 2 h after meals to avoid possible interference with postprandial hypotension [38]. Participants remained in a resting supine position on a bed for 5 min before postural change. After supine rest for 5 min, participants were passively moved from the supine to the sitting position within 30 s and maintained in the sitting position for 3 min with the assistance of an assessor [16, 18]. Moreover, participants were instructed not to assist with the maneuver during the test. The test was immediately terminated if a participant demonstrated severe symptoms such as presyncope, and the participant was returned to a supine position. Self-reported symptoms associated with orthostatic hypotension, such as dizziness, lightheadedness, or blurred vision, were recorded at the end of the test.

Blood pressure was measured on the left arm using an automated sphygmomanometer (HEM-907; Omron Co., Ltd., Kyoto, Japan). Systolic and diastolic blood pressure were measured in the supine position twice within 1 min after 5 min of rest.

After the postural change, blood pressure variables were measured in the upright position every minute for 3 min. Orthostatic hypotension was defined as a maximum reduction of ≥ 10 mmHg in systolic blood pressure or ≥ 5 mmHg in diastolic blood pressure during the test [18]. Supine hypertension was defined as systolic blood pressure of ≥ 140 mmHg or diastolic blood pressure of ≥ 90 mmHg in the supine position [12]. Seated hypertension was defined as systolic blood pressure of ≥ 140 mmHg or diastolic blood pressure of ≥ 90 mmHg at 3 min of sitting [1].

### Statistical analysis

The G power computer program version 3.1.9.2 (Heinrich Heine University, Dusseldorf, Germany) [39] was used for sample size calculation to detect blood pressure variable differences during the sit-up test between participants with and without orthostatic hypotension. We used an estimated effect size of 0.80 for the unpaired t-test, based on a study comparing blood pressure variables in supine and sitting positions between older adults with and without orthostatic hypotension [40]. The sample size was estimated to be 80, considering a statistical power of 0.80, an alpha level of 0.05, an expected prevalence of 22.2%, and an effect size of 0.80.

The normality of distribution for all continuous variables was assessed using the Shapiro–Wilk test. Blood pressure variables during the sit-up test were compared between the groups with and without orthostatic hypotension using two-way repeated-measures analysis of variance (ANOVA), with the group as the between-subject factor and time (supine and 1–3 min of sitting) as the within-subject factor. An unpaired t-test with Bonferroni correction was used to compare blood pressure variables between the groups at each time point. Additionally, we compared the prevalence of supine and seated hypertension between participants with and without orthostatic hypotension and that of seated hypertension between participants with and without supine hypertension using Fisher’s exact test.

To examine the associations between the three types of blood pressure dysregulation detected by the sit-up test and adverse health outcomes, we compared participant characteristics between those with and without orthostatic hypotension, supine hypertension, and seated hypertension. The unpaired t-test was used for continuous variables, and Fisher’s exact test was used for categorical variables. Participant characteristics included demographic, clinical, and geriatric outcomes. We conducted a subgroup analysis of participant characteristics with significant differences between those with and without orthostatic hypotension, comparing those with isolated orthostatic hypotension and those with co- existing orthostatic hypotension and supine and/or seated hypertension. Fisher’s exact test was used for categorical variables, and one-way ANOVA was used for continuous variables.

Subsequently, we performed multivariate analyses to examine the independent associations of orthostatic hypotension, supine hypertension, and seated hypertension with each participant characteristic variable. The presence of orthostatic hypotension, supine hypertension, and seated hypertension served as independent variables in these analyses, whereas participant characteristics served as dependent variables. Multiple regression analyses were conducted with continuous dependent variables, whereas logistic regression analyses were performed with categorical dependent variables. Statistical analyses were performed using GraphPad Prism version 9.00 for Windows (GraphPad Software, San Diego, California, USA). Statistical significance was set at two-sided p < 0.05.

## Results

### Participants

The sit-up tests were conducted in 16 of the 24 classes owing to scheduling constraints, with 139 individuals attending these sessions. Among those, 114 underwent the sit-up test. Moreover, 12 individuals were excluded from the analysis because they were below 65 years of age, resulting in a final sample of 102 participants for analysis. Table 1 lists the participant characteristics. Of all participants, 12 (11.8%) had at least one missing value in clinical and geriatric outcomes (Additional File 1). The final sample had a mean age of 75.0 ± 6.0 years, with 67 females (65.7%).

**Table 1.**
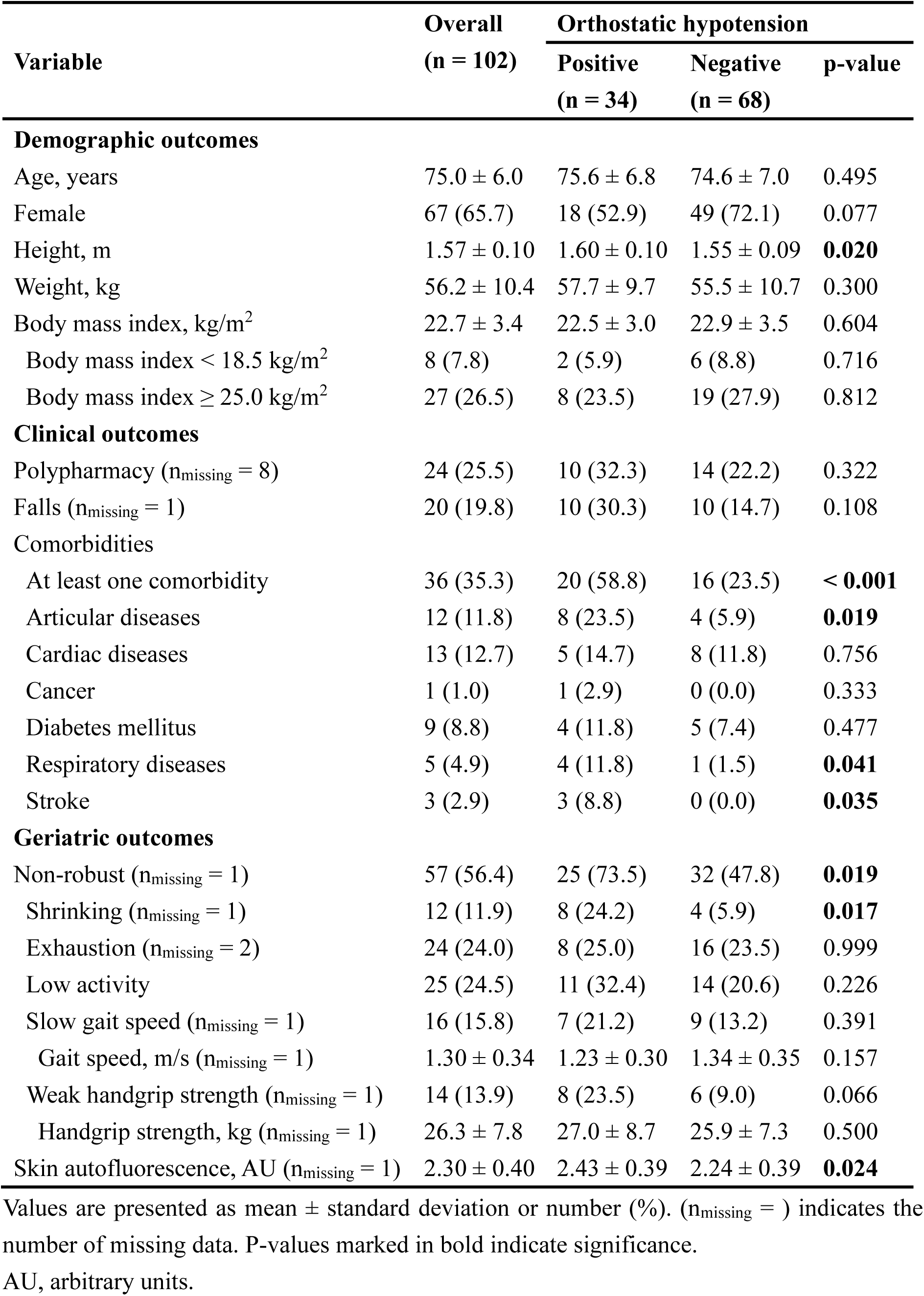
Comparisons of participant characteristics between those with and without orthostatic hypotension.

| Variable | Overall<br>(n = 102) | Orthostatic hypotension |  |  |
| --- | --- | --- | --- | --- |
|  |  | Positive<br>(n = 34) | Negative<br>(n = 68) | p-value |
| Demographic outcomes |  |  |  |  |
| Age, years | 75.0 ± 6.0 | 75.6 ± 6.8 | 74.6 ± 7.0 | 0.495 |
| Female | 67 (65.7) | 18 (52.9) | 49 (72.1) | 0.077 |
| Height, m | 1.57 ± 0.10 | 1.60 ± 0.10 | 1.55 ± 0.09 | **0.020** |
| Weight, kg | 56.2 ± 10.4 | 57.7 ± 9.7 | 55.5 ± 10.7 | 0.300 |
| Body mass index, kg/m<sup>2</sup> | 22.7 ± 3.4 | 22.5 ± 3.0 | 22.9 ± 3.5 | 0.604 |
| Body mass index < 18.5 kg/m<sup>2</sup> | 8 (7.8) | 2 (5.9) | 6 (8.8) | 0.716 |
| Body mass index ≥ 25.0 kg/m<sup>2</sup> | 27 (26.5) | 8 (23.5) | 19 (27.9) | 0.812 |
| Clinical outcomes |  |  |  |  |
| Polypharmacy (n<sub>missing</sub> = 8) | 24 (25.5) | 10 (32.3) | 14 (22.2) | 0.322 |
| Falls (n<sub>missing</sub> = 1) | 20 (19.8) | 10 (30.3) | 10 (14.7) | 0.108 |
| Comorbidities |  |  |  |  |
| At least one comorbidity | 36 (35.3) | 20 (58.8) | 16 (23.5) | **< 0.001** |
| Articular diseases | 12 (11.8) | 8 (23.5) | 4 (5.9) | **0.019** |
| Cardiac diseases | 13 (12.7) | 5 (14.7) | 8 (11.8) | 0.756 |
| Cancer | 1 (1.0) | 1 (2.9) | 0 (0.0) | 0.333 |
| Diabetes mellitus | 9 (8.8) | 4 (11.8) | 5 (7.4) | 0.477 |
| Respiratory diseases | 5 (4.9) | 4 (11.8) | 1 (1.5) | **0.041** |
| Stroke | 3 (2.9) | 3 (8.8) | 0 (0.0) | **0.035** |
| Geriatric outcomes |  |  |  |  |
| Non-robust (n<sub>missing</sub> = 1) | 57 (56.4) | 25 (73.5) | 32 (47.8) | **0.019** |
| Shrinking (n<sub>missing</sub> = 1) | 12 (11.9) | 8 (24.2) | 4 (5.9) | **0.017** |
| Exhaustion (n<sub>missing</sub> = 2) | 24 (24.0) | 8 (25.0) | 16 (23.5) | 0.999 |
| Low activity | 25 (24.5) | 11 (32.4) | 14 (20.6) | 0.226 |
| Slow gait speed (n<sub>missing</sub> = 1) | 16 (15.8) | 7 (21.2) | 9 (13.2) | 0.391 |
| Gait speed, m/s (n<sub>missing</sub> = 1) | 1.30 ± 0.34 | 1.23 ± 0.30 | 1.34 ± 0.35 | 0.157 |
| Weak handgrip strength (n<sub>missing</sub> = 1) | 14 (13.9) | 8 (23.5) | 6 (9.0) | 0.066 |
| Handgrip strength, kg (n<sub>missing</sub> = 1) | 26.3 ± 7.8 | 27.0 ± 8.7 | 25.9 ± 7.3 | 0.500 |
| Skin autofluorescence, AU (n<sub>missing</sub> = 1) | 2.30 ± 0.40 | 2.43 ± 0.39 | 2.24 ± 0.39 | **0.024** |

Values are presented as mean ± standard deviation or number (%). (n<sub>missing</sub> = ) indicates the number of missing data. P-values marked in bold indicate significance.
AU, arbitrary units.

Additionally, 36 participants (35.3%) had at least one comorbidity, and 57 (56.4%) were classified as non-robust. Among those classified as non-robust, 50 were pre-frail.

### Comparisons of blood pressure variables during the sit-up test between participants with and without orthostatic hypotension

Although all participants were asymptomatic during the sit-up test, a total of 34 participants (33.3%) met either the systolic or diastolic blood pressure criteria for orthostatic hypotension. Specifically, 26 showed systolic orthostatic hypotension, whereas two exhibited diastolic orthostatic hypotension. Six participants with orthostatic hypotension met the systolic and diastolic criteria for orthostatic hypotension.

Figure 1 shows blood pressure variables during the sit-up test in participants with and without orthostatic hypotension. The two-way repeated-measures ANOVA revealed significant interactions between group and time for systolic (F_(3,297)_ = 47.0, p < 0.001; Figure 1A) and diastolic (F_(3,297)_ = 26.5, p < 0.001; Figure 1B) blood pressure, indicating that participants with orthostatic hypotension showed a greater decrease in systolic blood pressure and a smaller increase in diastolic blood pressure after standing than those without orthostatic hypotension. Participants with orthostatic hypotension showed significantly higher mean values of systolic blood pressure in the supine position than those without orthostatic hypotension (t = 3.363, p = 0.005), whereas no significant differences were observed between the groups at any time point during the sitting period (Figure 1A). Participants with orthostatic hypotension demonstrated significantly lower mean values of diastolic blood pressure at 2 min of sitting than those without orthostatic hypotension, whereas no significant differences were observed between the groups at other time points (Figure 1B).

**Figure 1.**
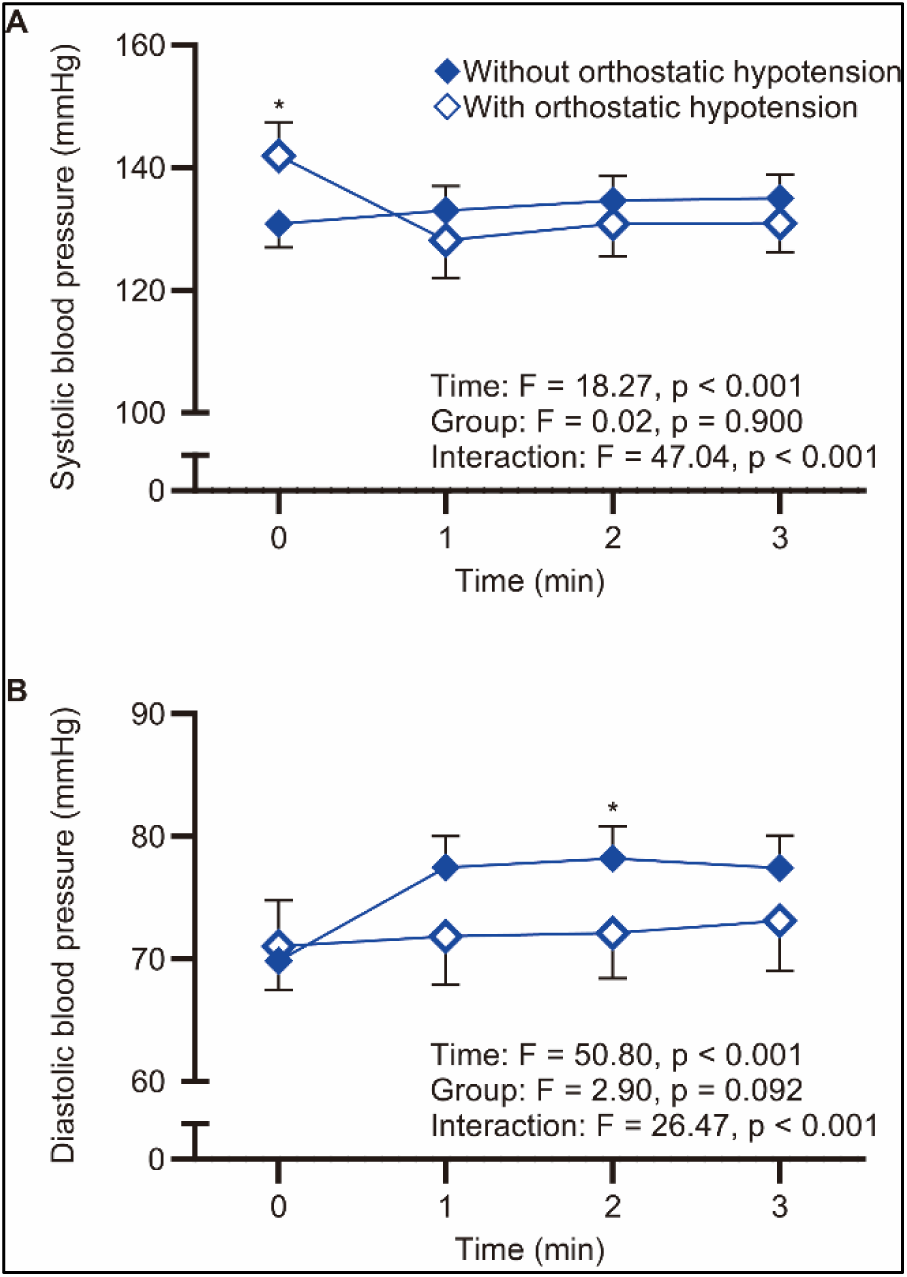
Blood pressure variables during the sit-up test in participants with and without orthostatic hypotension. (A) Systolic and (B) diastolic blood pressure changes during the sit-up test. The white and blue diamonds represent the mean blood pressure values in the groups with and without orthostatic hypotension, respectively, at each time point. The error bars indicate 95% confidence intervals. The x-axis represents the time after the postural change; therefore, data at x = 0 correspond to data in the supine position. The asterisks indicate significant differences between the groups with and without orthostatic hypotension (p < 0.05, Bonferroni multiple comparison test).

Overall, 36 participants (35.3%) exhibited supine hypertension, whereas 33 (32.4%) had seated hypertension. Additional File 2 describes the blood pressure dysregulation status of participants. Of the 34 participants with orthostatic hypotension, nine had both supine and seated hypertension, nine had supine hypertension alone, and 16 had isolated orthostatic hypotension. A significantly higher proportion of participants with orthostatic hypotension (52.9%, n = 18) had supine hypertension compared to those without orthostatic hypotension (26.5%, n = 18, p = 0.015), although no significant difference was observed in the proportion of participants with seated hypertension between those with (26.5%, n = 9) and without (35.3%, n = 24, p = 0.501) orthostatic hypotension. Additionally, a significantly larger proportion of participants with supine hypertension (63.9%, n = 23) had seated hypertension compared to those without supine hypertension (15.2%, n = 10, p < 0.001).

### Comparisons of participant characteristics between those with and without orthostatic hypotension

Table 1 shows the results of the comparative analysis. Regarding demographic outcomes, the mean height values were significantly higher in participants with orthostatic hypotension than those without (t = 2.356, p = 0.020). The remaining demographic outcomes did not significantly differ between the groups (p > 0.05).

Regarding clinical outcomes, a greater proportion of participants with orthostatic hypotension (58.8%, n = 20) self-reported at least one comorbidity compared to those without orthostatic hypotension (23.5%, n = 16, p < 0.001). Specifically, participants with orthostatic hypotension demonstrated higher prevalences of articular diseases (23.5%, n = 8 vs. 5.9%, n = 4, p = 0.019), respiratory diseases (11.8%, n = 4 vs. 1.5%, n = 1, p = 0.041), and stroke (8.8%, n = 3 vs. 0.0%, n = 0, p = 0.035) compared to participants without orthostatic hypotension.

Regarding geriatric outcomes, a greater proportion of participants with orthostatic hypotension (73.5%, n = 25) was classified as non-robust compared to those without orthostatic hypotension (47.8%, n = 32, p = 0.019). Shrinking prevalence was significantly higher in participants with orthostatic hypotension (24.2%, n = 8) than in those without (5.9%, n = 4, p = 0.017). Furthermore, mean skin autofluorescence values were significantly higher in participants with orthostatic hypotension than in those without (t = 2.300, p = 0.024).

Additionally, Additional File 3 shows the results of subgroup analysis in participants with orthostatic hypotension. No significant differences were observed in any of the variables among participants with isolated orthostatic hypotension, those with both orthostatic hypotension and supine hypertension, and those with all three types of blood pressure dysregulation (p > 0.05).

### Comparisons of participant characteristics according to supine hypertension and seated hypertension status

Table 2 illustrates the results of comparisons of participant characteristics according to supine hypertension and seated hypertension status. Participants with supine hypertension were significantly older than those without supine hypertension (t = 2.508, p = 0.014). Additionally, a larger proportion of participants with supine hypertension (40.6%, n = 13) self-reported polypharmacy compared to those without supine hypertension (17.7%, n = 11, p = 0.024). However, no participant characteristics showed a significant difference between those with and without seated hypertension (p > 0.05).

**Table 2.** Comparisons of participant characteristics according to supine hypertension and seated hypertension status.

| Variable | Supine hypertension |  |  | Seated hypertension |  |  |
| --- | --- | --- | --- | --- | --- | --- |
|  | Positive<br>(n = 36) | Negative<br>(n = 66) | p-value | Positive<br>(n = 33) | Negative<br>(n = 69) | p-value |
| **Demographic outcomes** |  |  |  |  |  |  |
| Age, years | 77.2 ± 6.2 | 73.7 ± 7.0 | **0.014** | 75.6 ± 5.0 | 74.6 ± 7.7 | 0.492 |
| Female | 23 (63.9) | 44 (66.7) | 0.829 | 25 (75.8) | 42 (60.9) | 0.182 |
| Height, m | 1.58 ± 0.09 | 1.57 ± 0.10 | 0.567 | 1.56 ± 0.09 | 1.58 ± 0.10 | 0.433 |
| Weight, kg | 57.6 ± 9.8 | 55.5 ± 10.7 | 0.324 | 56.1 ± 10.1 | 56.3 ± 10.6 | 0.931 |
| Body mass index, kg/m<sup>2</sup> | 23.1 ± 3.6 | 22.5 ± 3.3 | 0.376 | 23.0 ± 3.4 | 22.6 ± 3.3 | 0.590 |
| Body mass index < 18.5 kg/m<sup>2</sup> | 2 (5.6) | 6 (9.1) | 0.709 | 1 (3.0) | 7 (10.1) | 0.432 |
| Body mass index ≥ 25.0 kg/m<sup>2</sup> | 13 (36.1) | 14 (21.2) | 0.158 | 10 (30.3) | 17 (24.6) | 0.633 |
| **Clinical outcomes** |  |  |  |  |  |  |
| Polypharmacy (n<sub>missing</sub> = 8) | 13 (40.6) | 11 (17.7) | **0.024** | 11 (35.5) | 13 (20.6) | 0.137 |
| Falls (n<sub>missing</sub> = 1) | 10 (28.6) | 10 (15.2) | 0.122 | 7 (21.2) | 13 (19.1) | 0.796 |
| **Comorbidities** |  |  |  |  |  |  |
| At least one comorbidity | 16 (44.4) | 20 (30.3) | 0.194 | 12 (36.4) | 24 (34.8) | 0.999 |
| Articular diseases | 5 (13.9) | 7 (10.6) | 0.750 | 4 (12.1) | 8 (11.6) | 0.999 |
| Cardiac diseases | 7 (19.4) | 6 (9.1) | 0.212 | 7 (21.2) | 6 (8.7) | 0.111 |
| Cancer | 1 (2.8) | 0 (0.0) | 0.353 | 0 (0.0) | 1 (1.4) | 0.999 |
| Diabetes mellitus | 3 (8.3) | 6 (9.1) | 0.999 | 2 (6.1) | 7 (10.1) | 0.714 |
| Respiratory diseases | 3 (8.3) | 2 (3.0) | 0.342 | 1 (3.0) | 4 (5.8) | 0.999 |
| Stroke | 1 (2.8) | 2 (3.0) | 0.999 | 0 (0.0) | 3 (4.3) | 0.549 |
| **Geriatric outcomes** |  |  |  |  |  |  |
| Non-robust (n<sub>missing</sub> = 1) | 23 (65.7) | 34 (51.5) | 0.208 | 20 (62.5) | 37 (53.6) | 0.518 |
| Shrinking (n<sub>missing</sub> = 1) | 5 (14.3) | 7 (10.6) | 0.748 | 2 (6.1) | 10 (14.7) | 0.328 |
| Exhaustion (n<sub>missing</sub> = 2) | 11 (32.4) | 13 (19.7) | 0.217 | 10 (30.3) | 14 (20.9) | 0.327 |
| Low activity | 8 (22.2) | 17 (25.8) | 0.811 | 6 (18.2) | 19 (27.5) | 0.338 |
| Gait speed, m/s (n<sub>missing</sub> = 1) | 1.36 ± 0.40 | 1.27 ± 0.29 | 0.187 | 1.38 ± 0.40 | 1.27 ± 0.30 | 0.123 |
| Slow gait speed (n<sub>missing</sub> = 1) | 7 (19.4) | 9 (13.6) | 0.570 | 4 (12.1) | 12 (17.4) | 0.573 |
| Handgrip strength, kg (n<sub>missing</sub> = 1) | 26.9 ± 8.4 | 25.9 ± 7.5 | 0.535 | 25.4 ± 8.0 | 26.7 ± 7.7 | 0.440 |
| Weak handgrip strength (n<sub>missing</sub> = 1) | 5 (14.3) | 9 (13.6) | 0.999 | 4 (12.5) | 10 (14.5) | 0.999 |
| Skin autofluorescence, AU (n<sub>missing</sub> = 1) | 2.36 ± 0.44 | 2.27 ± 0.38 | 0.285 | 2.25 ± 0.39 | 2.33 ± 0.40 | 0.359 |

Values are presented as mean ± standard deviation or number (%). (n<sub>missing</sub> = ) indicates the number of missing data. P-values marked in bold indicate significance.
AU, arbitrary units.

### Independent associations of orthostatic hypotension, supine hypertension, and seated hypertension with participant characteristics

Table 3 shows the results of the multiple and logistic regression analyses. Orthostatic hypotension was significantly associated with a higher height [partial regression coefficient = 0.04, 95% confidence interval (CI) = 0.00–0.08, p = 0.048], higher prevalences of at least one comorbidity (odds ratio = 4.50, 95%CI = 1.74–11.6, p = 0.002) and articular diseases (odds ratio = 5.68, 95%CI = 1.40–23.10, p = 0.015), and a higher proportion of non-robust participants (odds ratio = 3.08, 95%CI = 1.17–8.08, p = 0.022) and those with shrinking (odds ratio = 4.32, 95%CI = 1.11–16.80, p = 0.032), even when controlling for supine and seated hypertension status.

**Table 3.** Independent associations of three types of blood pressure dysregulation with participant characteristics.

| Variable | Orthostatic hypotension |  | Supine hypertension |  | Seated hypertension |  |
| --- | --- | --- | --- | --- | --- | --- |
|  | Coefficient<br>(95%CI) | p-value | Coefficient<br>(95%CI) | p-value | Coefficient<br>(95%CI) | p-value |
| **Demographic outcomes** |  |  |  |  |  |  |
| Age* | B = -0.18<br>(-3.22, 2.87) | 0.909 | B = 4.08<br>(0.63, 7.53) | **0.021** | B = -1.08<br>(-4.49, 2.34) | 0.533 |
| Female | OR = 0.50<br>(0.20, 1.26) | 0.144 | OR = 0.71<br>(0.24, 2.13) | 0.538 | OR = 2.31<br>(0.74, 7.21) | 0.149 |
| Height* | B = 0.04<br>(0.00, 0.08) | **0.048** | B = 0.01<br>(-0.04, 0.06) | 0.726 | B = -0.02<br>(-0.06, 0.03) | 0.492 |
| Weight* | B = 1.54<br>(-3.13, 6.20) | 0.514 | B = 2.34<br>(-2.95, 7.62) | 0.382 | B = -1.24<br>(-6.48, 3.99) | 0.638 |
| Body mass index* | B = -0.59<br>(-2.11, 0.93) | 0.442 | B = 0.81<br>(-0.91, 2.53) | 0.352 | B = -0.06<br>(-2.11, 0.93) | 0.927 |
| Body mass index < 18.5 kg/m<sup>2</sup> | OR = 0.55<br>(0.09, 3.24) | 0.510 | OR = 1.24<br>(0.18, 8.66) | 0.826 | OR = 0.24<br>(0.02, 2.56) | 0.235 |
| Body mass index ≥ 25.0 kg/m<sup>2</sup> | OR = 0.58<br>(0.20, 1.65) | 0.305 | OR = 2.80<br>(0.90, 8.73) | 0.076 | OR = 0.75<br>(0.24, 2.32) | 0.618 |
| **Clinical outcomes** |  |  |  |  |  |  |
| Polypharmacy | OR = 1.40<br>(0.50, 3.93) | 0.528 | OR = 2.59<br>(0.78, 8.61) | 0.119 | OR = 1.28<br>(0.39, 4.26) | 0.684 |
| Falls | OR = 2.08 | 0.179 | OR = 2.09 | 0.249 | OR = 0.82 | 0.757 |
|  | (0.71, 6.07) |  | (0.60, 7.32) |  | (0.23, 2.95) |  |
| **Comorbidities** |  |  |  |  |  |  |
| At least one comorbidity | OR = 4.50<br>(1.74, 11.6) | **0.002** | OR = 1.21<br>(0.40, 3.61) | 0.737 | OR = 1.15<br>(0.38, 3.48) | 0.810 |
| Articular diseases | OR = 5.68<br>(1.40, 23.10) | **0.015** | OR = 0.69<br>(0.14, 3.44) | 0.651 | OR = 1.55<br>(0.30, 7.92) | 0.598 |
| Cardiac diseases | OR = 1.28<br>(0.34, 4.81) | 0.720 | OR = 1.48<br>(0.34, 6.45) | 0.605 | OR = 2.38<br>(0.57, 9.98) | 0.236 |
| Cancer | NA | NA | NA | NA | NA | NA |
| Diabetes mellitus | OR = 1.61<br>(0.36, 7.14) | 0.531 | OR = 1.01<br>(0.17, 5.88) | 0.992 | OR = 0.59<br>(0.09, 3.98) | 0.591 |
| Respiratory diseases | OR = 6.26<br>(0.61, 64.80) | 0.124 | OR = 2.85<br>(0.34, 24.1) | 0.336 | OR = 0.34<br>(0.03, 4.22) | 0.403 |
| Stroke | NA | NA | NA | NA | NA | NA |
| **Geriatric outcomes** |  |  |  |  |  |  |
| Non-robust | OR = 3.08<br>(1.17, 8.08) | **0.022** | OR = 1.12<br>(0.39, 3.21) | 0.826 | OR = 1.53<br>(0.54, 4.32) | 0.426 |
| Shrinking | OR = 4.32<br>(1.11, 16.80) | **0.035** | OR = 1.63<br>(0.35, 7.59) | 0.534 | OR = 0.31<br>(0.05, 1.97) | 0.212 |
| Exhaustion | OR = 0.95<br>(0.33, 2.71) | 0.927 | OR = 1.79<br>(0.55, 5.80) | 0.331 | OR = 1.20<br>(0.38, 3.82) | 0.756 |
| Low activity | OR = 1.89<br>(0.69, 5.16) | 0.216 | OR = 0.83<br>(0.25, 2.73) | 0.755 | OR = 0.68<br>(0.20, 2.30) | 0.535 |
| Slow gait speed | OR = 1.37 | 0.597 | OR = 2.20 | 0.252 | OR = 0.43 | 0.252 |
|  | (0.43, 4.41) |  | (0.57, 8.52) |  | (0.10, 1.84) |  |
| Gait speed* | B = -0.12 | 0.109 | B = 0.10 | 0.259 | B = 0.05 | 0.560 |
|  | (-0.27, 0.03) |  | (-0.07, 0.27) |  | (-0.12, 0.22) |  |
| Weak handgrip strength | OR = 3.51 | 0.051 | OR = 0.69 | 0.625 | OR = 1.14 | 0.864 |
|  | (1.00, 12.30) |  | (0.15, 3.08) |  | (0.25, 5.20) |  |
| Handgrip strength* | B = 0.39 | 0.827 | B = 1.99 | 0.326 | B = -2.26 | 0.259 |
|  | (-3.14, 3.92) |  | (-2.01, 5.99) |  | (-6.21, 1.69) |  |
| Skin autofluorescence* | B = 0.15 | 0.093 | B = 0.11 | 0.279 | B = -0.12 | 0.233 |
|  | (-0.03, 0.33) |  | (-0.09, 0.31) |  | (-0.32, 0.08) |  |

The asterisks in the variable column indicate that multiple regression analysis was employed as a multivariate analysis owing to the continuous nature of the dependent variable. P-values marked in bold indicate significance.
B, partial regression coefficient; CI, confidence interval; NA, not applicable; OR, odds ratio.

Supine hypertension was significantly associated with older age (partial regression coefficient = 4.08, 95%CI = 0.63–7.53, p = 0.021) after adjusting for orthostatic hypotension and seated hypertension status. No significant associations were observed between seated hypertension and participant characteristics (p > 0.05).

## Discussion

To the best of our knowledge, this study is the first to investigate the associations of orthostatic hypotension detected by the sit-up test with blood pressure variables during the test and poor health conditions in community-dwelling older adults. Participants with orthostatic hypotension demonstrated a greater decrease in systolic blood pressure, a smaller increase in diastolic blood pressure, and higher supine systolic blood pressure during the sit-up test compared to those without orthostatic hypotension. Consequently, more than 50% of participants with orthostatic hypotension showed supine hypertension. Moreover, orthostatic hypotension showed independent associations with adverse health outcomes, regardless of supine and seated hypertension status. Our findings contribute valuable insights for the application of the sit-up test in preventive health screenings for older adults.

### Differences in blood pressure variables during the sit-up test between participants with and without orthostatic hypotension

The prevalence of orthostatic hypotension in our study (33.3%) was higher than that reported in previous studies using conventional standing tests (22.2%) [11], which may be attributed to our methodological approach in diagnosing orthostatic hypotension. Consensus guidelines recommend that a reduction in systolic blood pressure of 30 mmHg, rather than the standard 20 mmHg, during conventional standing tests is a more appropriate diagnostic criterion for orthostatic hypotension in individuals with supine hypertension [3]. However, such diagnostic thresholds have not been established for the sit-up test. Therefore, we applied the same criteria regardless of supine hypertension status, which likely contributed to the higher prevalence of orthostatic hypotension in this study compared to previous studies [11].

Participants with orthostatic hypotension demonstrated significantly higher supine systolic blood pressure than those without orthostatic hypotension, in line with previous studies [40–43], resulting in a higher prevalence of supine hypertension. This relationship may be explained by several physiological mechanisms, such as age-related physiological impairments in baroreflex sensitivity and autonomic cardiovascular regulation [2, 12, 44]. Conversely, the prevalence of seated hypertension did not significantly differ between those with and without orthostatic hypotension. This result may be attributed to two distinct blood pressure responses to the sit-up test. First, participants with orthostatic hypotension exhibited a greater orthostatic reduction in systolic blood pressure, resulting in no significant difference in seated systolic blood pressure between those with and without orthostatic hypotension. Second, those with orthostatic hypotension demonstrated a blunted orthostatic increase in diastolic blood pressure, despite no significant difference in supine diastolic blood pressure between participants with and without orthostatic hypotension.

Diastolic blood pressure typically increases by 5–10 mmHg upon standing owing to peripheral vasoconstriction and stroke volume reduction [45]. Therefore, the blunted orthostatic increase in diastolic blood pressure likely reflects impaired arterial baroreflex-mediated function, which is considered the primary hemodynamic mechanism underlying orthostatic hypotension in older adults [42, 46].

### Associations of orthostatic hypotension detected by the sit-up test with adverse health outcomes

Our findings revealed that orthostatic hypotension was significantly associated with higher proportions of participants with at least one comorbidity and those classified as non-robust, even after adjusting for supine and seated hypertension. In contrast, seated hypertension showed no significant associations with any of the demographic, clinical, and geriatric outcomes. These findings suggest that the sit-up test can provide more valuable information for blood pressure management in older adults compared to conventional seated blood pressure measurement.

Previous studies of older adults have documented associations of orthostatic hypotension with age-related diseases and physical frailty [20, 47–52]. Orthostatic hypotension remains the most common measurement of autonomic dysfunction [53]. Autonomic dysfunction is associated with various age-related disorders, such as articular diseases, cardiac diseases, cancer, diabetes, respiratory diseases, and stroke [54–59]. Furthermore, older adults with physical frailty are more likely to have autonomic dysfunction [53]. Therefore, autonomic dysfunction may underlie the observed associations of orthostatic hypotension with poor health conditions and non-robust status.

The observed association between orthostatic hypotension and non-robust status may support the longitudinal associations of orthostatic hypotension with mortality and morbidity reported in previous studies [3–10], considering the association between physical frailty and an increased risk of future adverse health outcomes [25, 26]. Additional longitudinal studies are required to determine whether orthostatic hypotension detected by the sit-up test has stronger associations with future adverse events compared to supine and seated hypertension.

### Study limitations

This study has several limitations that warrant consideration. First, the cross-sectional design precludes establishing causality between blood pressure dysregulation and health outcomes.

Longitudinal studies are required to determine whether orthostatic hypotension precedes adverse health conditions. Second, recruitment from community health promotion classes may have introduced a selection bias toward health-conscious individuals. Third, our study included only independently ambulatory older adults, thus excluding those who truly require the sit-up test. Future studies should include older adults who cannot independently stand or are at high risk of falling while standing.

Finally, clinical outcomes, including the number of prescribed medications, history of falls within a year, and comorbidities, were based on self-reported data, which may be subject to recall and reporting biases.

## Conclusion

Orthostatic hypotension was associated with a greater decrease in systolic blood pressure, a smaller increase in diastolic blood pressure, and higher supine systolic blood pressure during the sit-up test in community-dwelling older adults. Therefore, more than 50% of participants with orthostatic hypotension had supine hypertension. In addition, orthostatic hypotension detected by the sit-up test was associated with adverse health outcomes independently of supine and seated hypertension. Therefore, the application of the sit-up test in routine health screenings may enhance the early identification and management of health deterioration in community-dwelling older adults.

## Data Availability

The datasets used and/or analyzed during the current study are available from the corresponding author on reasonable request.

## List of abbreviations

AGEs: advanced glycation end products
ANOVA: analysis of variance
CI: confidence interval

## Declarations

### Ethics approval and consent to participate

This study protocol was approved by the appropriate ethics committee of Shinshu University (approval number: 6281). All participants provided written informed consent before enrolment in the study.

### Consent for publication

Not applicable.

### Authors’ contributions

Conceptualization: KO, YY; Data curation: KO, YY; Formal analysis: KO; Funding acquisition: KO; Investigation: KO, YY; Methodology: KO, YY; Project administration: YY; Resources: KO, YY; Software: KO; Supervision: YY; Validation: KO; Visualization: KO; Witting-original draft: KO; Writing-review and editing: KO, YY; Approval of final manuscript: KO, YY.

## Acknowledgments

We would like to thank the staff at the Shiga Ward Community Development Center for their help and support. We also appreciate Editage (www.editage.com) for English language editing.

## Additional File 1

**Supplementary Table 1.** Summary of missing values in clinical and geriatric outcomes.

| Variable | Value |
| --- | --- |
| **Clinical outcomes** |  |
| Polypharmacy | 8 (7.8) |
| Falls | 1 (1.0) |
| **Geriatric outcomes** |  |
| Non-robust | 1 (1.0) |
| Shrinking | 1 (1.0) |
| Exhaustion | 2 (2.0) |
| Slow gait speed | 1 (1.0) |
| Gait speed | 1 (1.0) |
| Weak handgrip strength | 1 (1.0) |
| Handgrip strength | 1 (1.0) |
| Skin autofluorescence | 1 (1.0) |

Values are presented as numbers (%).

## Additional File 2

**Supplementary Table 2.**
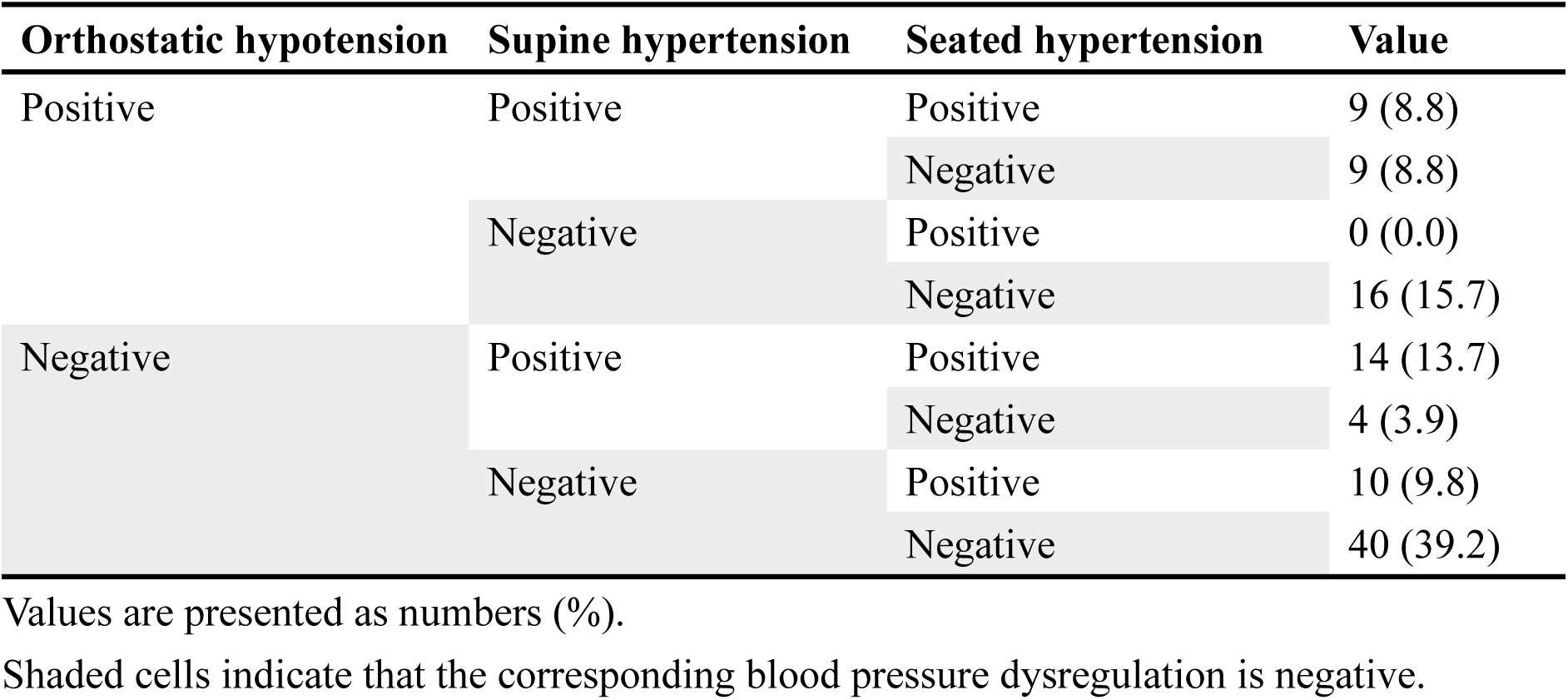
Blood pressure dysregulation status among participants.

## Additional File 3

**Supplementary Table 3.**
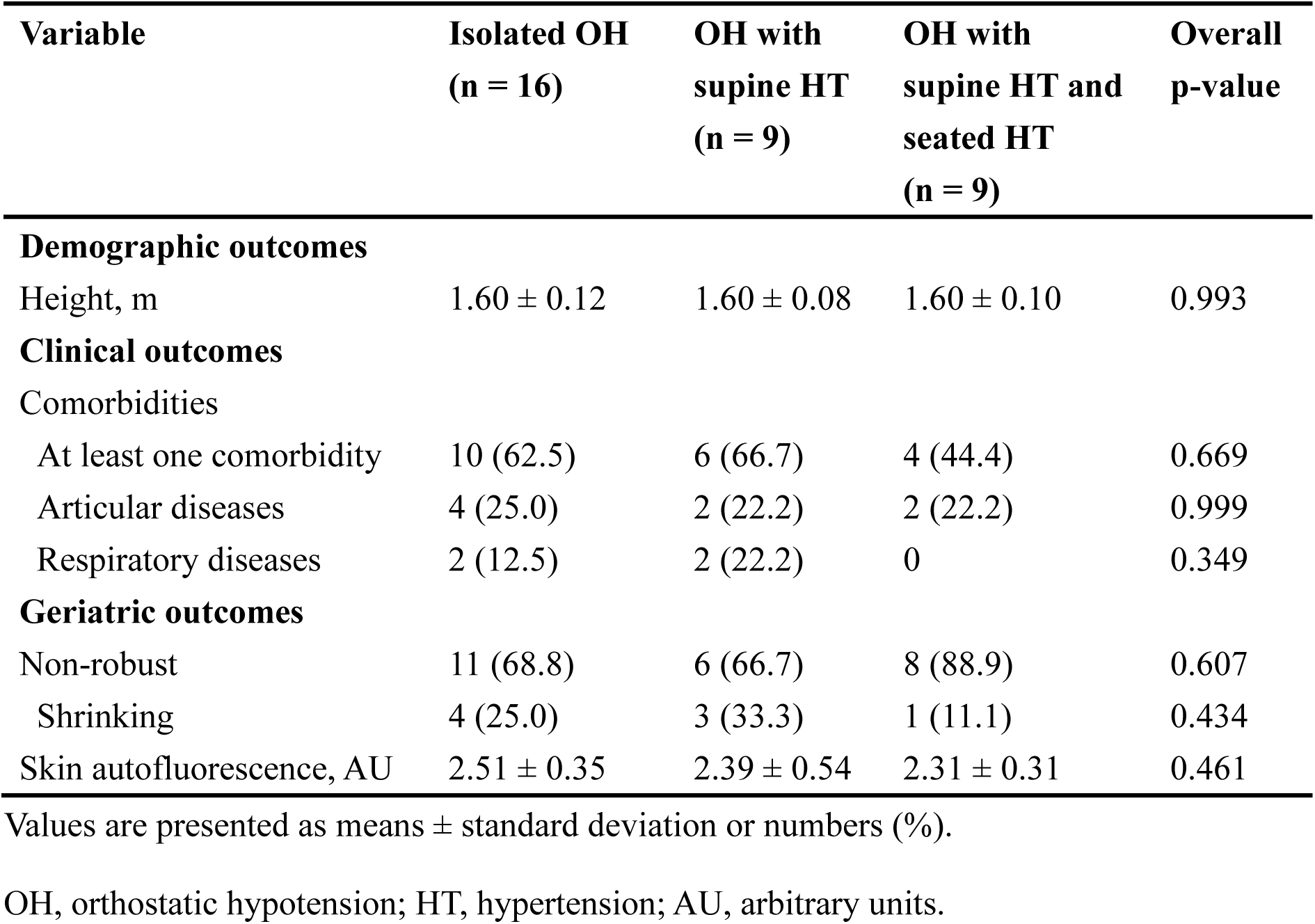
Subgroup analyses of participants with orthostatic hypotension.

| Variable | Isolated OH<br>(n = 16) | OH with<br>supine HT<br>(n = 9) | OH with<br>supine HT and<br>seated HT<br>(n = 9) | Overall<br>p-value |
| --- | --- | --- | --- | --- |
| **Demographic outcomes** |  |  |  |  |
| Height, m | 1.60 ± 0.12 | 1.60 ± 0.08 | 1.60 ± 0.10 | 0.993 |
| **Clinical outcomes** |  |  |  |  |
| Comorbidities |  |  |  |  |
| At least one comorbidity | 10 (62.5) | 6 (66.7) | 4 (44.4) | 0.669 |
| Articular diseases | 4 (25.0) | 2 (22.2) | 2 (22.2) | 0.999 |
| Respiratory diseases | 2 (12.5) | 2 (22.2) | 0 | 0.349 |
| **Geriatric outcomes** |  |  |  |  |
| Non-robust | 11 (68.8) | 6 (66.7) | 8 (88.9) | 0.607 |
| Shrinking | 4 (25.0) | 3 (33.3) | 1 (11.1) | 0.434 |
| Skin autofluorescence, AU | 2.51 ± 0.35 | 2.39 ± 0.54 | 2.31 ± 0.31 | 0.461 |

Values are presented as means ± standard deviation or numbers (%).
OH, orthostatic hypotension; HT, hypertension; AU, arbitrary units.

